# “The transcriptome-wide association search for genes and genetic variants which associate with BMI and gestational weight gain in women with type 1 diabetes”

**DOI:** 10.1101/2020.06.26.20137265

**Authors:** Agnieszka H. Ludwig-Słomczyńska, Michał T. Seweryn, Przemysław Kapusta, Ewelina Pitera, Urszula Mantaj, Katarzyna Cyganek, Paweł Gutaj, Łucja Dobrucka, Ewa Wender-OŻegowska, Maciej T. Małecki, Paweł P. Wołkow

## Abstract

**Background:** Clinical data suggest that BMI and gestational weight gain (GWG) are strongly interconnected phenotypes, however the genetic basis of the latter is rather unclear. Here we aim to find genes and genetic variants which influence BMI and/or GWG.

**Methods:** We have genotyped 316 type 1 diabetics using Illumina Infinium Omni Express Exome-8 v1.4 arrays. The GIANT, ARIC and T2D-GENES summary statistics were used for TWAS (performed with PrediXcan) in adipose tissue. Next, the analysis of association of imputed expression with BMI in the general and diabetic cohorts (Analysis 1 and 2) or GWG (Analysis 3 and 4) was performed, followed by variant association analysis (1Mb around identified loci) with the mentioned phenotypes.

**Results:** In Analysis 1 we have found 175 BMI associated genes and 19 variants (p<10^−4^) which influenced GWG, with the strongest association for rs11465293 in *CCL24* (p=3.18E-06). Analysis 2, with diabetes included in the model, led to discovery of 1812 BMI associated loci and 207 variants (p<10^−4^) influencing GWG, with the strongest association for rs9690213 in *PODXL* (p=9.86E-07). In Analysis 3, among 648 GWG associated loci, 2091 variants were associated with BMI (FDR<0.05). In Analysis 4, 7 variants in GWG associated loci influenced BMI in the ARIC cohort.

**Conclusions:** Here, we have shown that loci influencing BMI might have an impact on GWG and GWG associated loci might influence BMI, both in the general and T1DM cohorts. The results suggest that both phenotypes are related to insulin signaling, glucose homeostasis, mitochondrial metabolism, ubiquitinoylation and inflammatory responses.

## INTRODUCTION

In the recent years, a significant attention was paid to transcriptomic-wide association studies (TWAS), which enable the prediction of gene-level associations between the ‘imputed expression based on genetic data’ and complex phenotypes (Gamazon et al., 2015). This is possible, since it was shown that a proportion of GWAS risk variants co-localize with genetic variants, which regulate gene expression (i.e. expression quantitative trait loci, eQTL) (Hormozdiari et al., 2016). The prediction of gene expression based on genotype removes the noise created by environmental factors as well as the potential reverse causation (when the trait affects gene expression), therefore the TWAS analysis not only increases the statistical power, but also enables to focus on regions for which finding a functional interpretation of the associations is often easier (B. Li et al., 2018; Mancuso et al., 2017).

Gestational weight gain (GWG) has been extensively studied over the last several years as it was suggested that it’s inadequacy may lead to adverse both maternal (preeclampsia, hypertension, obesity later in life, cesarean section) and neonatal (preterm birth, stillbirth, inadequate neonatal weight – small or large for gestational age, obesity later in life) outcomes (*Influ. Pregnancy Weight Matern. Child Heal*., 2007; Kominiarek & Peaceman, 2017; Voerman et al., 2019). Simultaneously, clinicians tried to optimize pregnancy care for women with type 1 diabetes whose pregnancy outcomes (including developmental abnormalities, spontaneous abortions, neonatal hyperglycemia, neonatal hyperinsulinemia, maternal retinopathy, maternal nephropathy, preeclampsia) are far worse than in women from the general population (Celia et al., 2016). For a long time it was believed that it stemmed from maternal hyperglycemia, however, even though glycemic goals for these women have been achieved (HbA1c < 6.0%) still large for gestational age (LGA) or macrosomic neonates are born more frequent than in general population (Bashir, Naem, Taha, Konje, & Abou-Samra, 2019; Dori-Dayan et al., 2020; Scifres, Feghali, Althouse, Caritis, & Catov, 2014). Thus, there must be other potential contributors to adverse maternal and fetal outcomes (Mastella et al., 2018; Rys, Ludwig-Slomczynska, Cyganek, & Malecki, 2018; Secher et al., 2014). Among them are also those which affect the general population - maternal body lipids, pre-pregnancy BMI and GWG (McWhorter et al., 2018).

Even though GWG can have an impact on the health of future generations, to date, most of the performed studies are retrospective or observational and their main goal was to assess the relationship between GWG and environmental factors (Nunnery, Ammerman, & Dharod, 2018; Siega-Riz, Bodnar, Stotland, & Stang, 2020), while the genetic risks for inadequate GWG have been scarcely analyzed. In a recent paper, the authors show that 43% and 26% of the variation in GWG can be explained by genetic factors in the first pregnancy and second pregnancy, respectively (Andersson et al., 2015). The first GWAS study on GWG in the general, multiethnic cohort was performed in 2018. The study has shown that 20% of the variability in GWG can be explained by maternal genetic variants. Unfortunately, it did not find significant associations of GWG with any genomic loci (Warrington et al., 2018).

Since, both environmental and genetic data show that there is a correlation between pre-pregnancy BMI and GWG (Luecke et al., 2018) we took the TWAS approach to study the genetic correlation between BMI and GWG - in particular, to find loci and variants which are associated with GWG and BMI, or (on the contrary) only with GWG. We find this particularly important, as the reports on the overlap of genes and/or variants which influence both phenotypes appear to be conflicting (Kawai, Nwosu, Kurnik, Harrell, & Stein, 2019; Lawlor et al., 2011).

In this study, we aimed to investigate the genetic factors associated with GWG from the perspective of the genetics of obesity. To this aim, we performed TWAS analyses on the GIANT cohort representing the general population and two diabetic cohorts - T2D-GENES and ARIC to find genes associated with BMI. The ARIC and T2D-GENES cohorts were used since diabetes mellitus itself might influence BMI and creates a special metabolic context, while no cohort with patients with type 1 diabetes was available. Only regions which were associated with BMI in these cohorts were subjected to the analysis of association with GWG in T1DM patients. Secondly, we also searched for reverse associations and checked whether genes in TWAS associated with GWG in T1DM cohort might affect BMI in ARIC or GIANT cohorts. If so, we decided to search for genes which imputed expression correlated with BMI in order to restrict the analysis of genetic association with GWG to these genes only and conversely, to check whether the genes which correlate with GWG (at the imputed expression level) do influence BMI. Thus, this work comprises 4 different analyses, as presented in Figure 1.

**Figure 1.**
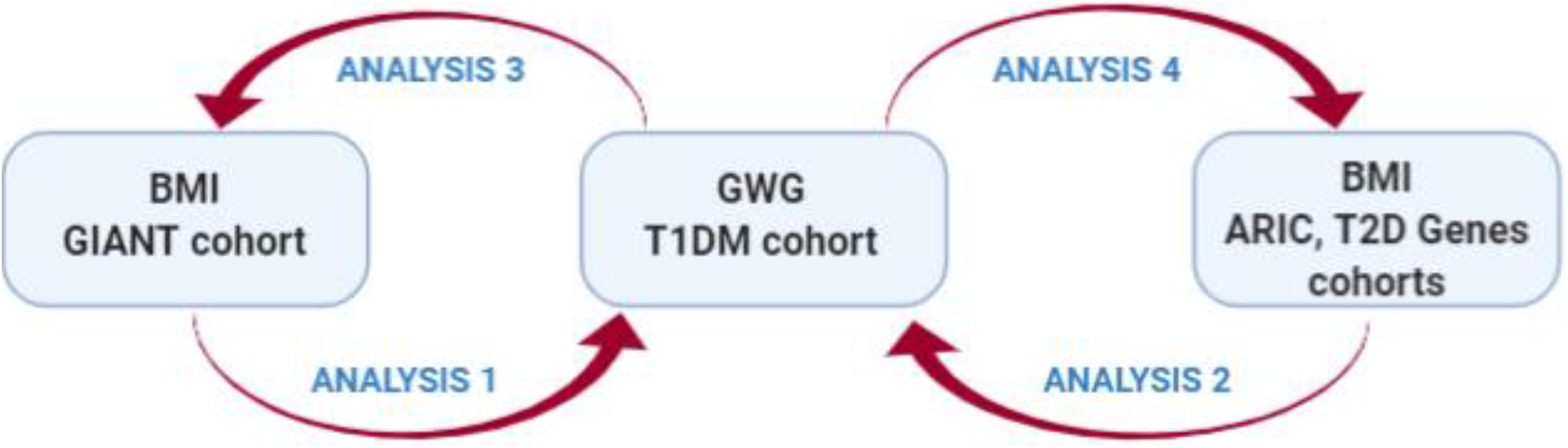
The scheme of the workflow. TWAS analyses to find genes associated with BMI in the general cohort (GIANT) and two diabetic cohorts (T2D-GENES, ARIC) as well as with GWG in T1DM cohort, followed by variant associations between the two phenotypes.

## METHODS

### Patients

Patients were recruited either in Department of Metabolic Diseases University Hospital in Krakow or in Division of Reproduction Department of Obstetrics, Gynecology and Gynecological Oncology, Poznan University of Medical Sciences. All patients enrolled in the study were women with type 1 diabetes (T1D) and treated with insulin. Pre-pregnancy weight was based either on women self-report or weight measurement (if the women were/was under antenatal care). Only singleton pregnancies were included. Pregnancies that resulted in a miscarriage or stillbirth were excluded. Total GWG was defined as the difference between last gestational weight before delivery and pre-pregnancy weight. Whole blood samples were drawn and stored at -80C. This study was approved by the Bioethical Committees of the Jagiellonian University and Poznan University of Medical Sciences and performed according to the Helsinki Declaration. Written informed consent was collected from all patients.

### Genotyping

DNA was extracted from whole blood with the use of automated nucleic acid extraction system Maxwell (Promega). Five hundred twenty-seven samples were genotyped on Illumina Infinium Omni Express Exome-8 v1.4 arrays. Only in term life births with information regarding age, pre-pregnancy BMI, GWG, diabetes duration, treatment method and daily insulin dose available were included in the final analysis.

### Data processing and imputation

The detailed protocol of the data processing, QC analysis and imputation is presented in (Ludwig-Slomczynska et al., 2018).

### Genotype Data and the analysis of genetic variants

The GWAS analysis on the T1DM cohort was performed on a group of 316 females with complete data on: age, parity, insulin dose prior pregnancy, pre-pregnancy BMI and GWG. The mixed-effects model approach was used as implemented in the package GENESIS in R. The random effect was associated with the individual ID and the genetic relatedness matrix was estimated via the PCARelate method in package GENESIS in R. The ‘null’ mixed effects model was considered as:

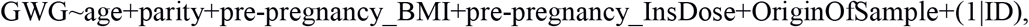

where the OriginOfSample is an indicator of the sample being collected in Division of Reproduction Department of Obstetrics, Gynecology and Gynecological Oncology, Poznan University of Medical Sciences. To the aim of testing significance of the (additive) effect of the genotype the Wald’s test statistics was used.

The summary statistics for BMI tested in the GIANT Consortium cohort were downloaded from https://portals.broadinstitute.org/collaboration/giant/images/1/15/SNP_gwas_mc_merge_nogc.tbl.uniq.gz

The genotype and phenotype data for the ARIC cohort (GENEVA study) were accessed via dbGaP. As far as the genotype data is concerned, the imputed data for participants of European ancestry were used as deposited under the phg000248.v1 code. Per-chromosome genotype probability data were transformed to dosages. From these dosages, genotype relatedness matrices were estimated on a per-chromosome basis and merged with the aid of the SNPRelate package in R. For the GWAS analysis the GENESIS package was used. As far as the phenotype files are concerned, these were accessed under the phs000090.v3 study code. The ‘null’ mixed effects model was defined as:

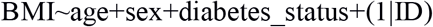

As GENEVA is a longitudinal study, for diabetic participants, we used the BMI and age measurement at the first time when the status of the participant was ‘diabetic’, whereas for the non-diabetic participants we used the measurements at the study entry.

The T2Dgenes genotype files were accessed via dbGaP under the phg000573.v1 code. The raw vcf files were transformed to dosages prior analysis. The phenotype data were accessed from the phs000462.v2 study code. Similarly to the data analysis in the ARIC cohort, we recorder BMI and age at the first time when the diabetic status was positive or at the study entry, otherwise.

The TWAS analysis was performed via the PrediXcan and MetaXcan software with Adipose as the tissue of interest. For the GIANT Cohort data, the MetaXcan framework was implemented based on the GTEx.v7 models. For the ARIC, T2Dgenes and T1DM cohorts the PrediXcan software was used also with GTEx.v7 models. In the TWAS analyses in the ARIC and T1DM cohorts the same independent variables were used as with the GWAS models defined above – with the exception that the classical linear models were used as implemented in the limma package (with no random effects). For the T2Dgenes TWAS analysis, from the ‘predicted expression’ matrix the principal components were first estimated and the first component was added to the models as it did not correlate with any independent variable – and it differ between families (as tested with the Kruskal-wallis test). As before the target analysis in the T2Dgenes cohort was performed with the aid of linear models as implemented in the limma package.

The FUMA analysis was performed through the webserver at fuma.ctglab.nl

The COJO and fast-BAT software were used as implemented by in the GTCA framework. The LD structure was estimated via the 1000 genomes data.

GO enrichment analysis was performed as implemented in the topGO package in R.

## RESULTS

### Gestational weight gain analysis in T1DM cohort

Our analysis comprised 316 women with T1DM for whom full phenotype data were available. The basic characterization of patients included in the analysis is presented in Table 1.

**Table 1.** The clinical characteristics of the T1DM cohort.

|  | Med; IQR |
| --- | --- |
| Age [years] | 29; (25.75, 32) |
| Pre-pregnancy BMI [kg/m2] | 23.41; (21.09, 26.2) |
| GWG [kg] | 14; (10.5, 18) |
| Diabetes duration [years] | 12; (6, 18) |
| **Treatment method – CSII [%]** | 0.55 |
| **Daily insulin dose before pregnancy [U]** | 38; (28, 50) |
| **Parity** | 1; (1, 2) |

PrediXcan was used to impute gene expression in subcutaneous and visceral adipose tissue based on imputed genotype data and the gene-based search for associations with GWG and BMI was performed using a linear model approach. Due to small cohort size, genes nominally significantly associated with GWG (442 genes in subcutaneous and 328 in visceral adipose tissue) and BMI (416 genes in subcutaneous and 326 in visceral adipose tissue) were considered for further analysis. It is worth noting that only sixty genes overlapped between the two phenotypes (Table S1). We further explored this phenomenon by looking at the strength of the association between gene expression for these two traits. We found a negative correlation between logFCs of genes associated with GWG and BMI. This is consistent with clinical recommendations as patients with high pregestational BMI are advised to restrict their GWG, however, we believe that the genetic background might also play a role.

### PrediXcan analysis on BMI in the general population (GIANT cohort) and variant associations with GWG in T1DM cohort - Analysis 1

Gene expression prediction in visceral and subcutaneous adipose tissue in the GIANT cohort (234069 patients, (Locke, Kahali, Berndt, Justice, & Pers, 2015)) as performed using PrediXcan software. One hundred seventy-five genes significantly associated (p < 1E-04) with BMI in the adipose (subcutaneous and visceral combined) tissue were found (Table S2). The Venn diagram of genes which affected BMI, GWG or both phenotypes is presented in Table S3. Only 15 genes influenced both phenotypes, 160 influenced only BMI, while 633 had an impact on GWG only (Figure 2).

**Figure 2.**
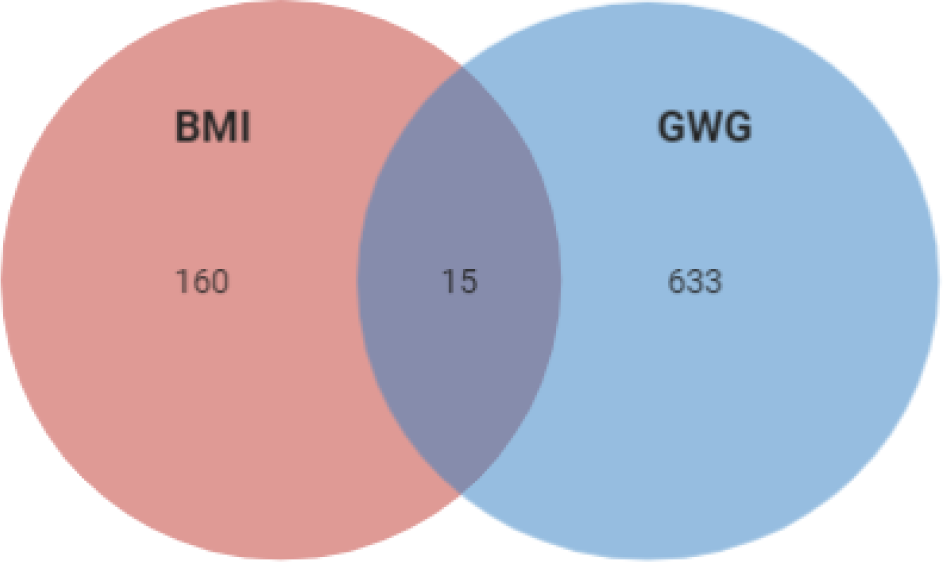
Venn diagram of the association of imputed gene expression which influence BMI in the Giant cohort, GWG in T1DM cohort and both phenotypes.

We further analyzed the loci of interest and searched for variants (within the 1Mb window around the gene) which influence BMI and/or GWG. We first mapped variants in 175 BMI associated genes (+/-500000 bp) and applied these restricted variant panels to the analysis of the association with GWG in the T1DM cohort. We filtered the results by MAF < 5% and p-value < 1E-04 and found an association with GWG for 19 variants in the T1DM cohort (Table 2).

**Table 2.** The list of variants localized to genes associated with BMI in the GIANT cohort which associate with GWG.

| SNP ID | chr | MAF [%] | B | p-value | Gene/ nearest gene | localization/type |
| --- | --- | --- | --- | --- | --- | --- |
| rs11465293 | 7 | 5.7 | 4.46 | 3.18E-06 | *CCL24* | missense |
| rs1978202 | 7 | 8.7 | 3.51 | 4.53E-06 | *CCL26* | intergenic |
| rs9347707 | 6 | 35.1 | 1.94 | 4.74E-05 | *PACRG* | intron |
| rs72849841 | 17 | 17.1 | -2.19 | 1.25E-04 | *RNF213* | missense |
| rs11807240 | 1 | 10.4 | 2.58 | 1.61E-04 | *FUBP1* | intron |
| rs9659938 | 1 | 10.4 | 2.58 | 1.61E-04 | *NEXN204* | intron |
| rs13340504 | 7 | 14.4 | 2.32 | 2.29E-04 | *CCL24* | intergenic |
| rs1541725 | 2 | 46.0 | 1.61 | 3.95E-04 | *AC009502.4* | intergenic |
| rs2136682 | 1 | 9.3 | 2.54 | 4.25E-04 | *GIPC2* | intron |
| rs4329088 | 6 | 46.2 | 1.63 | 4.33E-04 | *PACRG* | intron |
| rs886131 | 12 | 41.0 | -1.60 | 4.78E-04 | *RPL29P25* | noncoding<br>transcript |
| rs11037234 | 11 | 33.1 | 1.66 | 5.34E-04 | *RP11-111A24.2* | intergenic |
| rs8122282 | 20 | 45.7 | -1.49 | 6.44E-04 | *C20orf166* | intron |
| rs4808209 | 17 | 6.9 | 3.00 | 7.13E-04 | *ZNF101* | start loss |
| rs9352745 | 6 | 26.1 | 1.70 | 7.27E-04 | *LCA5* | intron |
| rs876373 | 12 | 43.4 | 1.50 | 7.29E-04 | *RPL29P25* | intron |
| rs6062267 | 20 | 45.3 | -1.49 | 7.94E-04 | *C20orf166* | intron |
| rs7961894 | 12 | 8.5 | 2.75 | 8.88E-04 | *WDR66* | intron |
| rs7564856 | 2 | 34.5 | -1.59 | 9.62E-04 | *SPEG* | intergenic |

The FUMA GWAS method further prioritized the variants obtained in the analysis. Twelve leading SNPs were detected. Three of them (rs7564856, rs11465293 and rs7961894) were associated with several traits in GWAS Catalog, mainly blood parameters, but, none significantly with metabolic outcome. At the same time however, these were also associated at the level of significance p-value < 1E-03 with LDL cholesterol levels, nonalcoholic fatty liver disease, body mass index, mean arterial pressure or metabolite levels. These variants were eQTLs for 40 genes in several tissues, while rs9659938, rs11807240, rs2136682 (*NEXN, NEXN-AS1*), rs7564856 (*SPEG*), rs9352745 (*SH3BGRL2*), rs13340504 (*CCL24*), rs11465293 (*CCL24*), rs886131 (*RPL29P25*) influenced gene expression in both subcutaneous and visceral adipose. Thus, we conclude that there is a subset of genes which influence both BMI and GWG as well as genetic variants associated with GWG which co-localize to these loci.

Next, we tried to determine, whether the signals for the two traits in the genes of interest are correlated, thus we checked whether the genetic variants associated with BMI and GWG are in linkage disequilibrium or they belong to different haplotypes. We performed the analysis of LD for all genes with variants significantly associated with GWG, however, below we present only the most interesting examples. In the gene *GPN3*, we have found 2 bins of eQTLs which are not in LD either with BMI or GWG variants. Variants associated with BMI also create a cluster, which is separate from a small cluster of 2 variants associated with GWG. One of GWG associated variants (rs876373) is in moderate LD with the BMI cluster (Figure S1a). In another gene, *PMS2P3*, we found a cluster of 3 variants which are associated with GWG, all in LD with one eQTL (rs707395). At the same time, variants associated with BMI form a separate, bin which is in moderate LD with a subset of eQTLs for this gene. This cluster is not in LD with the GWG bin or rs707395 (Figure S1b). The third example is the *STAG3L1* gene in which three clusters - GWG associated variants, BMI associated variants and eQTLs can be seen (r^2), however, in each, few members of any cluster are in LD with members of other clusters (D’) making them rather dependent on each other (Figure S1c).

### PrediXcan analysis on BMI in the diabetic cohorts (ARIC and T2D-GENES) and variant associations with GWG in T1DM cohort - Analysis 2

Since it is known that diabetes and its treatment might impact BMI, we searched for variants that might affect BMI in the context of diabetes status. Since no T1DM cohort was available, we used T2D patients’ cohorts – T2D-GENES (590 patients) and ARIC (8746 patients) (being aware that the two phenotypes obesity and diabetes are interconnected and T2D patients are predisposed to obesity, while T1DM patients not). After adjusting for diabetes, we found that 1812 genes were associated with BMI p < 0.05 (Table S4). Among them 135 influenced both phenotypes, 1677 influenced only BMI and 513 influenced GWG (Table S5, Figure 3).

**Figure 3.**
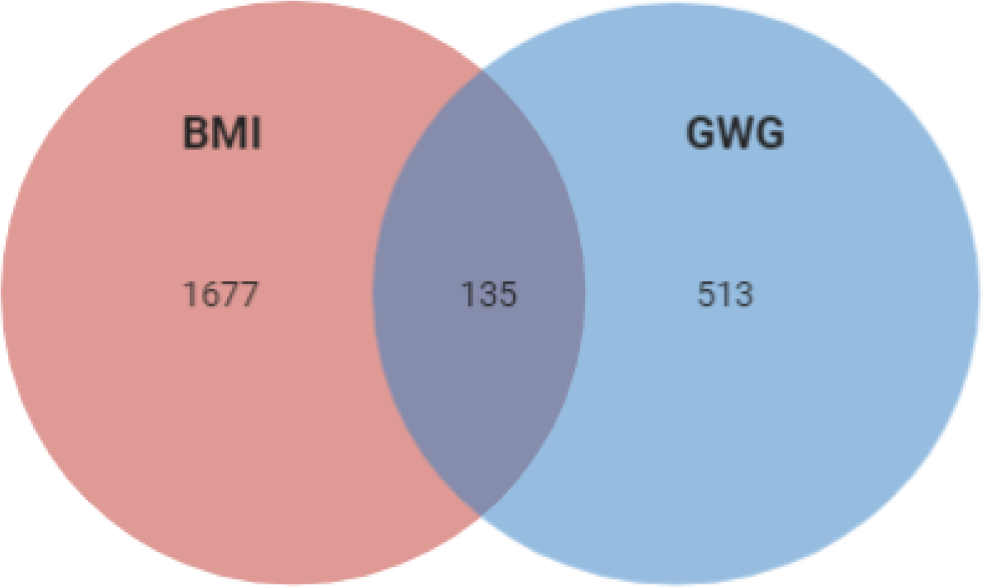
Overlap of the correlation of imputed expression of genes which influence BMI in ARIC and T2D-GENES cohorts, GWG in T1DM cohort and both phenotypes.

Again, we mapped variants to 1812 BMI associated genes (+/-500000bp) and used this panel to look for associations with GWG in T1DM cohort. When filtered by MAF < 5% and p-value < 1E-04 the analysis has shown that GWG correlates with 207 variants (Table S6). Table 3 shows 20 variants which were significant at the level of p-value < 1E-05.

**Table 3.** The list of variants localized to genes associated with BMI in T2D-GENES and ARIC cohorts which associate with GWG.

| SNP ID | chr | MAF [%] | B | p-value | Gene/ nearest gene | localization/type |
| --- | --- | --- | --- | --- | --- | --- |
| rs9690213 | 7 | 18.4 | 2.66 | 9.86E-07 | *PODXL* | regulatory region<br>variant |
| rs11465293 | 7 | 5.7 | 4.46 | 3.17E-06 | *CCL24* | missense |
| rs9393623 | 6 | 31.0 | 2.18 | 4.11E-06 | *CMAHP* | intron |
| rs4796675 | 17 | 22.2 | 2.38 | 4.40E-06 | *LINC00974* | noncoding transcript<br>exon variant |
| rs1978202 | 7 | 8.7 | 3.51 | 4.53E-06 | *CCL26* | intergenic |
| rs8080053 | 17 | 23.1 | 2.33 | 5.57E-06 | *LINC00974* | noncoding transcript<br>exon variant |
| rs12534221 | 7 | 17.7 | 2.55 | 5.85E-06 | *PODXL* | intergenic |
| rs6747327 | 2 | 35.6 | 2.04 | 1.35E-05 | *CAPN13* | intron |
| rs4742339 | 9 | 37.8 | 1.97 | 2.97E-05 | *KANK1* | intergenic |
| rs11714248 | 3 | 43.0 | -1.86 | 4.40E-05 | *TRANK1* | intron |
| rs9347707 | 6 | 35.1 | 1.94 | 4.74E-05 | *PACRG* | intron |
| rs1398095 | 3 | 25.6 | 2.15 | 4.85E-05 | *CAPDS* | intron |
| rs9940705 | 16 | 6.2 | 3.61 | 6.91E-05 | *FAM92B* | intron |
| rs2421052 | 5 | 43.1 | -1.86 | 7.63E-05 | *AC008691* | intron |
| rs6815557 | 4 | 39.2 | 1.82 | 8.24E-05 | *LOC107986225* | intergenic |
| rs12788347 | 11 | 10.8 | 2.92 | 8.72E-05 | *AL137224* | intron |
| rs11235519 | 11 | 24.2 | -2.02 | 8.93E-05 | *AP005019.4* | intergenic |
| rs6546217 | 2 | 11.7 | 2.64 | 9.89E-05 | *AC118345.1* | intergenic |
| rs7609526 | 2 | 11.1 | 2.68 | 9.97E-05 | *AC118345.1* | intergenic |

Most of the variants which influenced both GWG and BMI (p-value < 1E-05) were eQTLs associated with the expression of 13 genes (*LRRFIP2, MLH1, GOLGA4, CCL26, RHBDD2, AC004980, POR, CCL26, GTF2IRD2, UPK3B, ART2P, LINC00974, KTP15, KRT17*) in 14 tissues (Thyroid, Skeletal Muscle, Whole Blood, Artery, Cultured Fibroblasts, Tibial Nerve, Testis, Skin (Unexposed), Visceral Adipose Tissue, Minor Salivary Gland, Small Intestine, Colon, Brain (several regions).

FUMA analysis showed that among variants significant for association with GWG (p-value < 1E-04), 18 were listed for associations with multiple phenotypes in the GWAS Catalog. The KEGG analysis showed enrichment for autoimmunological disease systemic lupus erythematosus (adj. p-value = 3.32E-7) and taste transduction (adj. p-value = 2.18E-2). In the molecular functions GO enrichment analysis we found these variants to be responsible, among others, for carbohydrate binding (adj. p-value = 3.15E-8), bitter taste receptor activity (adj. p-value = 4.51E-5), trace amine receptor activity (adj. p-value = 9.69E-5) and taste receptor activity (adj. p-value = 9.04E-4) (FigureS2).

Finally, when comparing the lists of variants obtained in Analyses 1 and 2, we found a significant overlap, what supports our hypothesis that loci associated with BMI, both in the general population and diabetic cohorts may also associate with GWG. Several SNPs - rs7564856 (*SPEG*), rs1541725 (*AC009502*.*4*), rs9347707 (*PCARG*), rs4329088 (*PCARG*), rs1978202 (*CCL26*), rs13340504 (*CCL24*), rs11465293 (*CCL24*), rs11037234 (*RP11-111A24*.*2*), rs876373 (*RPL29P25*), rs886131 (*RPL29P25*), rs7961894 (*WDR66*), rs4808209 (*ZNF101*) were found to influence GWG in T1DM, ARIC, T2D-GENES and GIANT cohorts. We conclude, that despite the relatively large overlap between traits at the gene level, the genetic variants which localize with these loci most often are linked to a single phenotype only.

Next, we aimed to study the “reverse association” - i.e. we asked whether genes associated with GWG influence BMI as well.

### PrediXcan analysis on GWG in T1DM cohort and variant associations with BMI in the general population (GIANT cohort) - Analysis 3

Variants were mapped to 648 GWG associated genes (+/-500000 bp) (Table S1) and a search for associations with BMI in the GIANT cohort was performed. Among 394149 variants analyzed, 2091 were significantly associated with BMI (at FDR < 0.05). These significant SNPs encompassed 0.53% of the whole list of variants analyzed. In comparison, the significant SNPs in the analysis of BMI in the GIANT cohort make up 0.41%, what means that genes which influence GWG are significantly enriched in the association test for BMI.

We used the FUMA tool to prioritize these 2091 variants. Twelve lead SNPs and 12 genomic risk loci were found. Not surprisingly, the GWAS Catalog analysis has shown a significant enrichment for genes associated with BMI, body fat distribution, attendance to gym or sport groups, sleep duration, height, lipoprotein levels, low HDL-level cholesterol etc. as presented in Figure 4 and Table S7.

**Figure 4.**
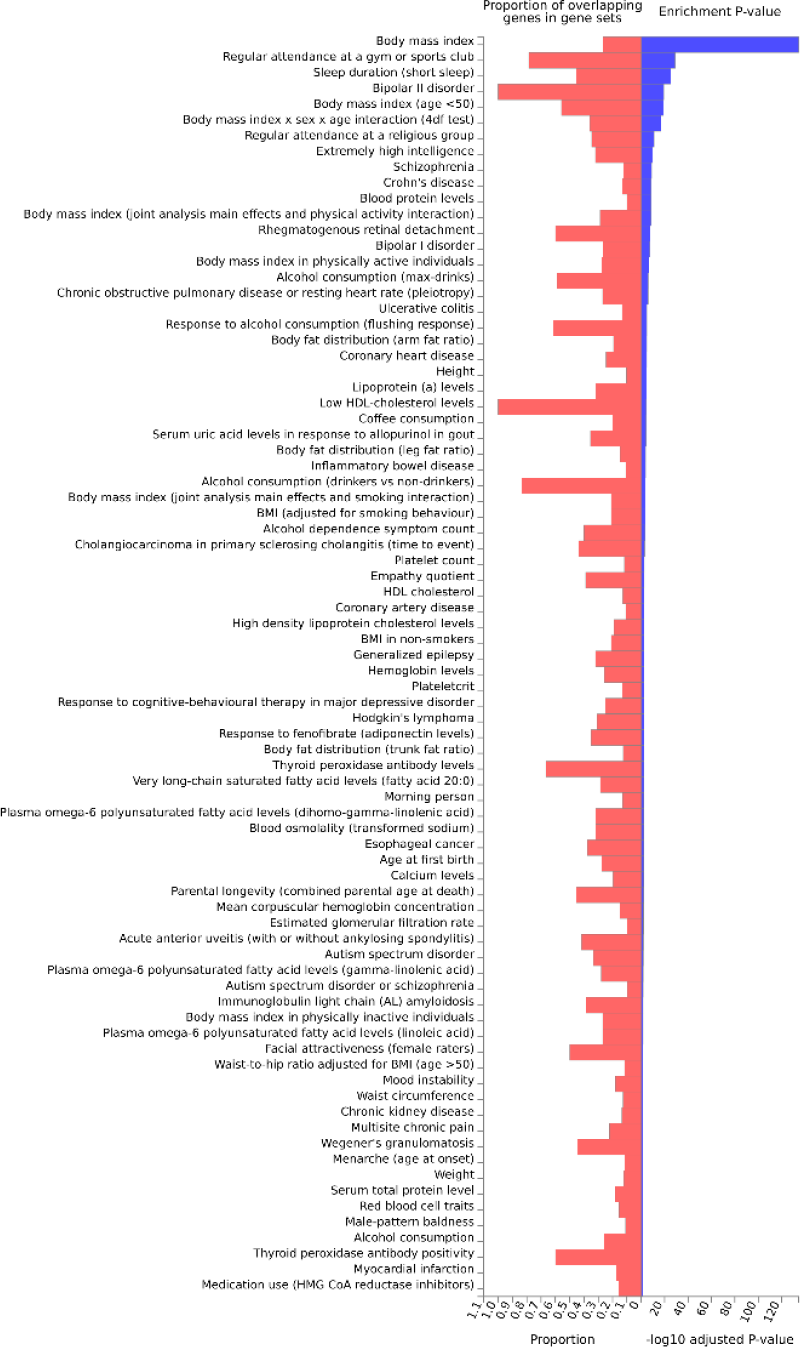
The enrichment analysis of variants localized to genes associated with GWG among those associated with BMI in the GIANT cohort.

Thus, we find a significant enrichment of variants associated with BMI in the general population within loci associated with GWG.

### PrediXcan analysis on GWG in T1DM cohort and variant associations with BMI in the diabetic cohort (ARIC) - Analysis 4

A GWAS analysis on BMI in the ARIC cohort was performed. Due to a large number of variants in the analysis we searched for lead SNPs using LD score regression. COJO analysis has returned 15 variants statistically significantly associated with the trait of interest (Table 4A).

**Table 4.** The list of variants localized to genes associated with BMI (A) and GWG (B) in ARIC cohort.

| SNP ID | chr | MAF [%] | B | p-value | Gene/ nearest gene | localization/type |
| --- | --- | --- | --- | --- | --- | --- |
| A. |  |  |  |  |  |  |
| rs138445632 | 2 | 7.92 | -2.88 | 4.54E-12 | AC021851,2 | intron |
| rs73646471 | 9 | 11.40 | -5.59 | 2.60E-10 | LOC105375974 | intergenic |
| rs72703233 | 9 | 8.47 | -1.57 | 1.31E-07 | DMRT2 | noncoding transcript |

|  |  |  |  |  |  | exon variant |
| --- | --- | --- | --- | --- | --- | --- |
| rs78231573 | 2 | 6.14 | -6.02 | 2.62E-07 | *LOC105373643* | intergenic |
| rs144768710 | 2 | 5.40 | 2.77 | 3.30E-07 | *CHN1* | intron |
| rs73419031 | 11 | 5.72 | -6.34 | 6.96E-07 | *SOX6* | intron |
| rs6456353 | 6 | 90.25 | 42.14 | 7.10E-07 | *E2F3* | intron |
| rs1486942 | 5 | 8.15 | -2.66 | 1.61E-06 | *LOC112267929* | intergenic |
| rs74036632 | 14 | 7.05 | -2.85 | 3.69E-06 | *NYNRIN* | 3'UTR |
| rs77346003 | 1 | 10.58 | -17.55 | 4.12E-06 | *ADIPOR1* | intergenic |
| rs73382735 | 9 | 5.68 | 4.49 | 4.28E-06 | *LOC105375955* | intergenic |
| rs12685009 | 9 | 8.42 | -8.94 | 7.74E-06 | *SLC24A2* | intergenic |
| rs34427781 | 22 | 19.46 | -2.54 | 8.47E-06 | *TCF20* | intron |
| rs13359900 | 5 | 15.98 | -27.84 | 8.95E-06 | *MCTP1* | intron |
| rs4131847 | 12 | 5.86 | 44.27 | 9.31E-06 | *CCDC60* | intergenic |
| B. |  |  |  |  |  |  |
| rs7875240 | 9 | 10.53 | -3.08 | 1.21E-06 | *PTPRD* | intron |
| rs1486942 | 5 | 8.15 | -2.66 | 1.61E-06 | *LOC112267929* | intergenic |
| rs74036632 | 14 | 7.05 | -2.85 | 3.69E-06 | *NYNRIN* | 3'UTR |
| rs34427781 | 22 | 19.46 | -2.54 | 8.47E-06 | *TCF20* | intron |

Only 4 variants from this list were in GWG associated genes in PrediXcan, three of which overlapped with those associated with BMI (Table 4B). None of the variants were reported in GWAS Catalog or PheWas for the association with any trait. 6 variants were eQTLs - two for genes in which they were localized rs13359900 and rs73419031 for *MCTP1* and *SOX6*, respectively; rs144768710 for *ATP5G3*, rs74036632 for *ZFHX2-AS1*, rs4131847 for *HSPB8*, while rs34427781 for several genes all localized 200000bp upstream or downstream from the variant.

Since the COJO analysis was performed on loci associated with GWG (+/-500000bp around the gene), it was possible that the most important variants influencing BMI were located outside those windows. Thus, we used a different tool, fastBAT, to find genes associated with BMI. The analysis revealed 8 genes which nominally associate with the trait of interest (Table 5).

**Table 5.** The list of variants localized to genes associated with BMI in ARIC cohort and associated with GWG as shown by fast-BAT.

| Gene/nearest gene | chr | localization | # of SNPs | p-value | Top SNP p-value | Top SNP |
| --- | --- | --- | --- | --- | --- | --- |
| *HLD-DQB* | 6 | intergenic | 4654 | 0.016 | 1.02E-06 | rs115258523 |
| *GATAD2* | 19 | intron | 1256 | 0.017 | 6.23E-05 | rs145702982 |
| *AC099791.3* | 1 | intron | 1800 | 0.020 | 8.85E-05 | rs2485748 |
| *LINC01141* | 1 | intron | 1501 | 0.027 | 9.95E-05 | rs12027661 |
| *HLA-DRB* | 6 | intergenic | 9354 | 0.029 | 1.89E-05 | rs139964305 |
| *HLA-B* | 6 | stop gained | 4017 | 0.044 | 2.11E-04 | rs149512147 |
| *LHFPL6* | 13 | intron | 1648 | 0.047 | 3.34E-06 | rs6563700 |

Again, none the of variants was previously mentioned to be significantly associated with any trait in GWAS Catalog or PheWas. Two were eQTLs - rs6563700 for *LHFPL6*, while rs12027661 for *CAMK2N1*.

## DISCUSSION

In this work we performed TWAS as well as single variant associations to determine the loci related to GWG and/or BMI. About 15% of loci overlap between BMI and GWG at the TWAS level. In the general population (GIANT cohort) the pathway analysis has shown that loci which contribute to both phenotypes affect mainly energy metabolism. Among the enriched pathways are regulation of mitochondrion (GO:1903749, GO:1903747, GO:0010822, GO:0070585, GO:0010821, GO:0006839) and Golgi apparatus (GO:0051683, GO:0048313, GO:0051645). There are also few pathways which affect transcription, polyadenylation or methylation of RNA (GO:1903311, GO:0016071, GO:1900363, GO:0080009). The analysis of the loci which TWAS associates with GWG only leads to necrosis (GO:0070266, GO:0097300, GO:0070265), protein kinases signaling (GO:0046330, GO:0043507, GO:0032874, GO:0070304, GO:0043506, GO:0007256) and metabolism of sugars (GO:0034033, GO:0034030, GO:0033866,GO:0009226). The BMI associated loci enrich cell cycle progression pathways (GO:0010972,GO:1902750) host to pathogen signaling (GO:0043921, GO:0052312, GO:0052472) and macromolecule metabolism (GO:0010604, GO:0034641, GO:0006139, GO:0009308, GO:0046483) and insulin signaling (GO:0046626, GO:1900076). The results of the GO enrichment analysis are presented in Supplementary Table S8 and Figures S3a, b.

The analysis of the overlap between BMI and GWG associated loci in the ARIC cohort and T1DM cohort respectively showed enrichment in the TGFβ signaling pathway (GO:0071559, GO:0071560), regulation of stem cell differentiation (GO:1901532, GO:1902036, GO:0060218, GO:2000736) and polyol pathway (GO:0019751, GO:0046173). Pathways associated with GWG were similar to those enriched in general population, however the important contribution of genes which affect antigen presentation could be seen (GO:0002495, GO:0002504, GO:0019886). The pathways characteristic for BMI associated loci were very much involved into response to environmental stimuli i.e. temperature (GO:0009266, GO:0009408), oxygen levels (GO:0070482, GO:0001666, GO:0036293) and sensory perception (GO:0050954, GO:0007600). The results of the GO enrichment analysis are presented in Supplementary Table S9 and Figures S4a, b.

At the genetic variant level, we detected SNPs in BMI associated loci which are related to GWG, and variants associated with both GWG and BMI (Analysis 1). At the same time, we performed a similar analysis with diabetes adjusted BMI and GWG, which led us to congruent findings (Analysis 2). These results point us towards inflammatory response, TGFβ signaling, ER stress and glucose homeostasis. Subsequently, we investigated the association of SNPs in loci which influenced GWG (based on TWAS) with BMI and found 2091 variants in the GIANT cohort (Analysis 3). A parallel analysis was done on diabetes adjusted BMI (in the ARIC cohort) – resulting in 15 variants localized to GWG associated loci (Analysis 4). The results of these analyses show an impact of lipid biosynthesis, appetite regulation, Ca^2+^ homeostasis (and ER stress) and inflammatory response on obesity. Our results point to the source of the genetic correlation between GWG and BMI and confirms the interconnection of the phenotypes. We compared the results of our gene focused analysis with GWAS on GWG published in 2018 (Warrington et al., 2018); even though variants from the discovery cohort did not replicate, their potential functional roles overlap with those found in our study. *TMEM163* is one of the best known genes associated with obesity, *LCORL* was shown to associate with height, *UGDH* with TGFβ signaling, *HLA-C* with autoimmune response, *HSD17B3* with fatty acids, *GLRX3* with oxidative stress, *RBM19* with ribosomal biogenesis, *SYT4* with Ca^2+^ binding and pancreatic functioning, *PSG5* with pregnancy development, while *NTS4* with neutrophilin signaling pathway. Our results are also congruent with those obtained in the computational search for functional annotation of 445 loci associated with obesity (Cheng et al., 2018). Therefore, we hypothesize that a further search for clinical phenotypes which affect GWG might allow identification of other loci that associate with it. Below we shed more light on the biological interpretation of our results and links between them (Fig. 5, 6).

**Figure 5.**
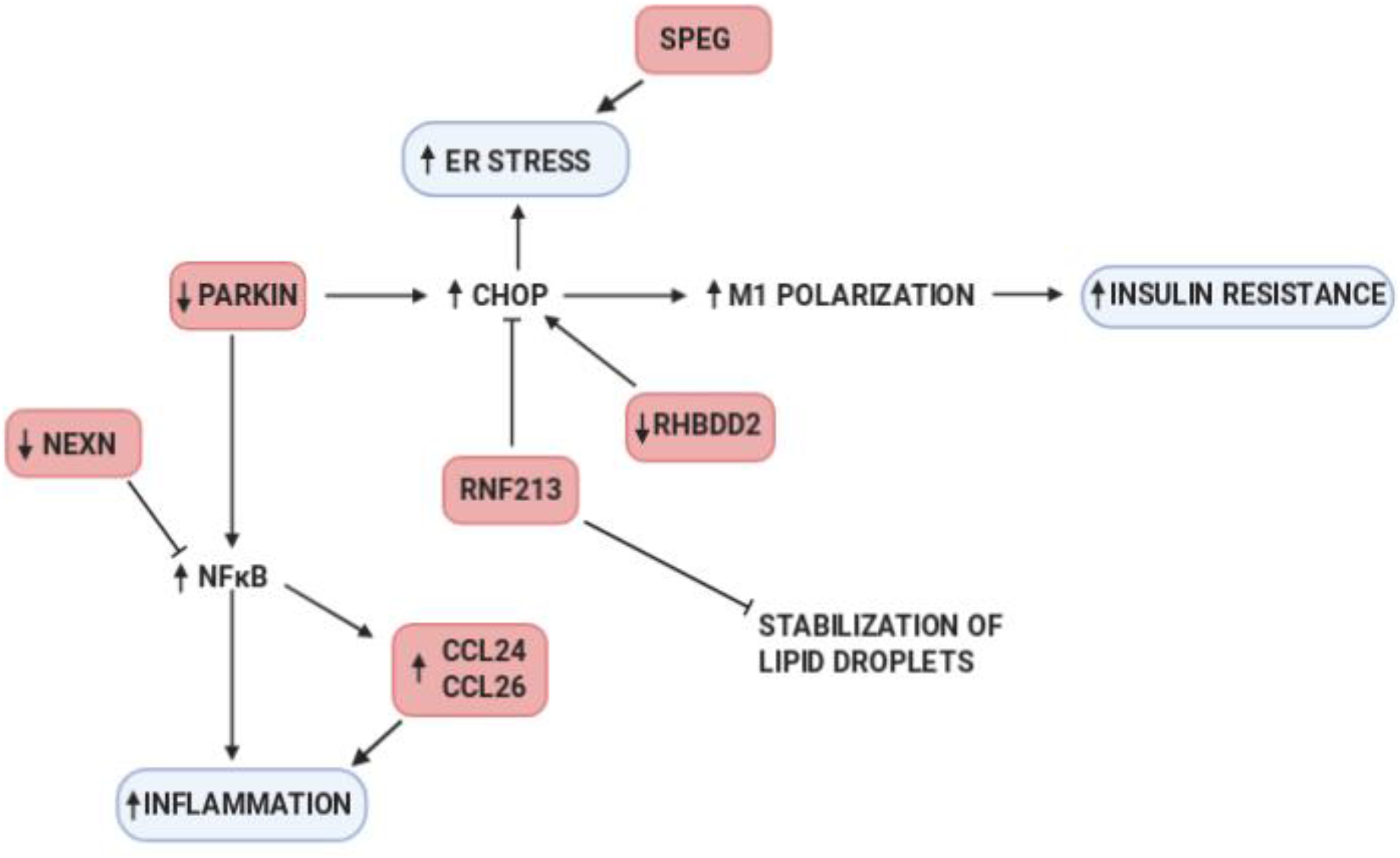
The schematic representation of pathways(blue) affected by genes which variants associate with GWG (pink).

**Figure 6.**
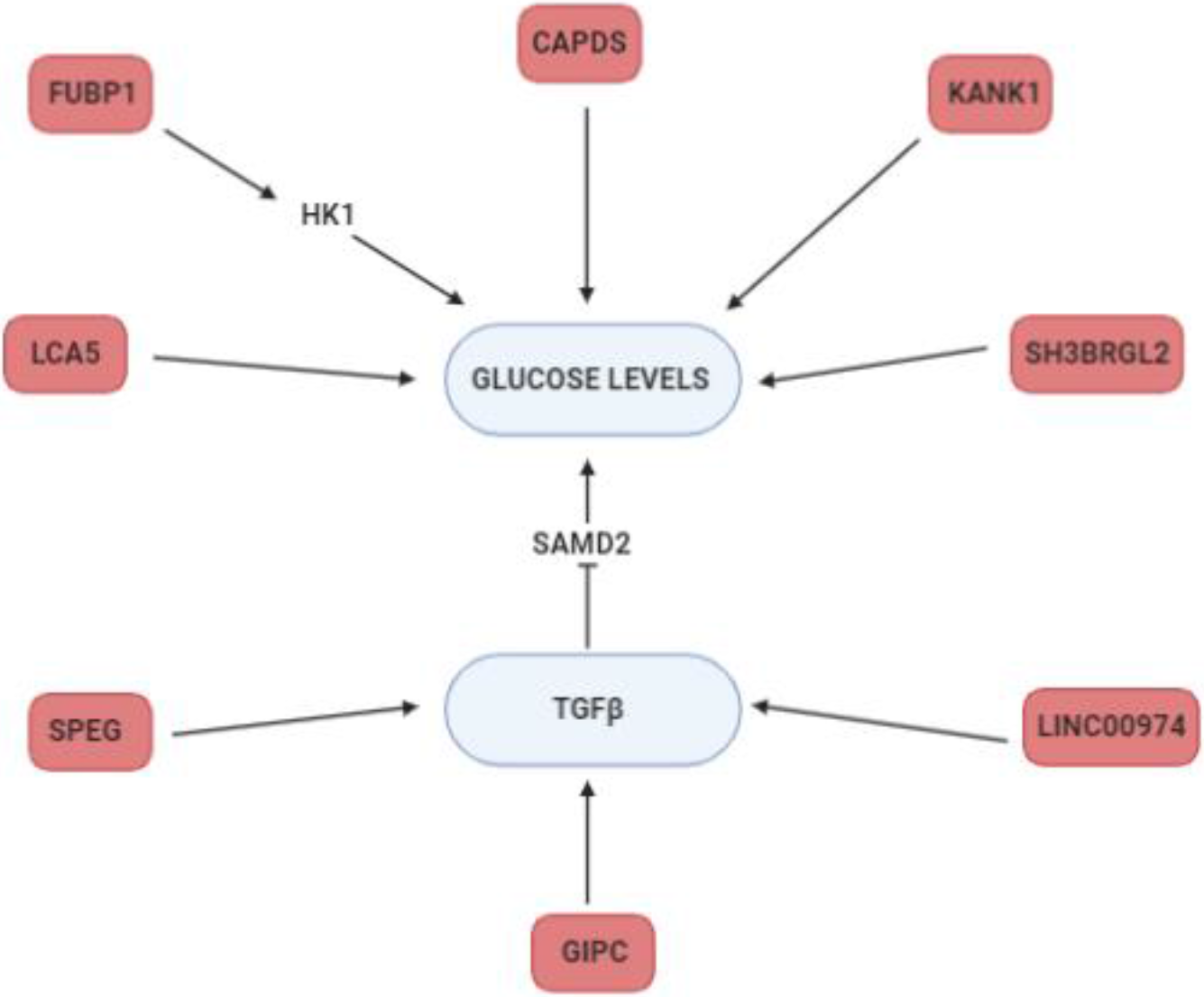
Genes which variants associate with GWG (pink) and that were reported to affect glucose levels or TGFβ signaling (blue).

In analyses 1 and 2, the strongest association with GWG was observed for SNPs in chemokine receptor ligands - rs1978202 – *CCL26* and rs13340504, rs11465293 - *CCL24*. Apart from variants located in these genes we identified several other (rs4742339, rs4796675, rs11235519) which serve as eQTLs for them. Chemokines are chemoattractant with the proinflammatory role. Their receptor expression was shown to correlate with BMI (being higher in obese individuals) (Ignacio, Gibbs, Lee, & Son, 2016) and BMI changes (decreased after bariatric surgery) (Gentili et al., 2016). CCL26 is also involved in adipose tissue beiging in response to cold (Finlin et al., 2017). At the same time, we find three variants rs9659938, rs11807240, rs2136682 associated with GWG in the *NEXN* gene (or serving as eQTL for it) also involved in the inflammatory response. Of note, the augmentation of NEXN antisense RNA (*NEXN-AS1*) inhibits TLR4 oligomerization and NFκB activity and leads to suppression of proinflammatory response (Hu et al., 2019). NEXN is also known for its abundant expression in striated muscles (Zhu et al., 2018) and cardiomyocytes; its expression is higher under high glucose conditions (Barbati et al., 2017) (Figure 5).

Interestingly, we find GWG associated variants in genes involved in TGFβ signaling: *SPEG, LINC000974, GIPC2*. The *GIPC2* gene (rs2136682 of which associates with GWG), belongs to the GIPC family known for regulating proliferation, cytokinesis or migration and involved in the trafficking of various transmembrane proteins. GIPC1 is necessary for cell-surface expression of transmembrane receptors, such as IGF1R and TGFβR3 (Katoh, 2013; Song et al., 2016). The inhibition of *LINC000974* leads to TGFβ secretion and repression of phosphorylated SMAD2 expression (Fang et al., 2019; Tang et al., 2014). Both TGFβ and SMADs are involved in insulin expression and signaling; SMAD4 deficiency was shown to improve glucose tolerance and glucose induced insulin release (Lin et al., 2009). Of note, three GWG associated variants (rs9690213, rs9393623, rs1978202) are eQTLs for *LINC000974* and four (rs8080053, rs1978202, rs4796675, rs8080053) are located in the gene itself (Figure 6).

The rs7564856, negatively associated with GWG is located in *SPEG*, and its expression is significantly greater in endurance than power athletes (Kusić et al., 2020). Also, at the protein level, SPEG differs between muscles of high-response vs low response rat trainers. The coimmunoprecipitation experiment with SPEG in the former showed a significant number of proteins involved in JNK and TGFβ signaling (Kusić et al., 2020), which, as said before, is known to influence insulin signaling and ER stress response (Z. Liu et al., 2019) (Figure 4, 5).

Two other variants - rs9347707 and rs4329088 - located in *PACRG1* gene, known to be involved in the insulin pathway. In *C. elegans* PACRG mutants have reduced insulin signaling. Also, PCARG plays a key role in the regulation of mitophagy (Stephenson et al., 2018) and ubiquitynylation (Loucks et al., 2016), its knockout leads to defective NFκB signaling (Meschede et al., 2020) and CHOP upregulation (Han et al., 2017), which in adipose tissue result in increased proinflammatory macrophage polarization (M1) and insulin resistance (Suzuki et al., 2017). Thus, we note that PACRG is also involved in ER stress. The *RHBDD2* gene is also involved in this biological process, with six eQTLs (rs4742339, rs11235519, rs1978202, rs13340504, rs4796675 and rs13340504) associated with GWG. It is worth noting that silencing of *RHBDD2* leads to increased expression of ATF6, IRE1, PERK, CRT, BiP, ATF4 and CHOP (*32*) (Figure 4).

The association of rs72849841 (in *RNF213*) with GWG draws our particular attention. Namely, in Akita mice (which develop diabetes spontaneously) the knockout of *RNF213* lowers glucose tolerance by 20% and leads to increased insulin contents in pancreases (by 150%) compared to wild type animals. This is obtained via the inhibition of ubiquitinoylation (RNF213 is a ubiquitin ligase) as shown by the 30% lower percentage of CHOP positive B-cells (Kobayashi et al., 2013). The depletion of *RNF213* protects cells from lipotoxicity (Piccolis et al., 2019), reduces palmitate-induced cell death and modifies non-mitochondrial oxygen consumption rate. Moreover, RNF213 is targeted to lipid droplets, which are specialized for neutral lipid storage and increases their abundance and stability in cells through the elimination of adipose triglyceride lipase (ATGL) (Sugihara et al., 2019). It is interesting to note, that the latter is also negatively regulated by insulin (Kershaw et al., 2006). *RNF213* variants in GWAS analyses suggestively associated (p-value ∼E-03) with diabetic polyneuropathy, portal hypertension, diabetic retinopathy and abnormal glucose (Figure 4). Its expression differed between cyclic and pregnant heifers (Forde et al., 2012).

We identified GWG associated loci involved in glucose homeostasis or pancreatic functioning (Figure 5) - e.g. rs11807240 in *FUBP1*, which upregulates the mRNA levels of the two hexokinase genes *Hk1* and *Hk2* - the rate limiting enzymes of glycolysis. A positive correlation between *FUBP1* mRNA and both of hexokinases was found in several types of cancers (Kang, Lee, Kim, Lee, & Kim, 2019). Variants in *LCA5* gene were shown to modify glucose response in a clinical trial of insulin and potassium (GIK) infusion in acute coronary syndromes (Ellis et al., 2015). The rs9352745 associated with GWG is an eQTL for *SH3BRGL2*, whose expression is associated with diabetes (type 1, type 2 and gestational) in a meta-analysis study. An interaction between *SH3BRGL2* SNP and total fat intake was found to affect LDL-PPD (Rudkowska et al., 2015). We identified GWG associated SNP rs7609526 in *KANK1* related to fasting proinsulin (Huyghe et al., 2013) and fasting plasma glucose (Hwang et al., 2015) in GWAS. *CADPS* (with GWG associated SNP rs1398095) plays a role in insulin secretion (Speidel et al., 2008). Cg14451730 located near *CADPS* is differentially methylated with HbA1c levels in the Taiwan Biobank cohort (Hsiung et al., 2019). The suggestive association with fasting plasma glucose of rs487321 in *CADPS* gene was found in Arabian population of T2D patients (Hebbar et al., 2020). We also found a variant correlating with GWG in *RPL29P25* gene. The presence of anti-60S ribosomal protein L29 (RPL29) antibody in human serum was shown to inhibit the proliferation of pancreatic cancer cells in various cancers and it is believed to be a novel candidate for a prognostic marker for unresectable pancreatic cancer (Muro, Miyake, Kato, Tsutsumi, & Yamamoto, 2015). The knockdown of *RPL29* leads to suppression of cell proliferation, induces cell arrest (at G0/G1phase), enhances cell apoptosis and decreases intracellular ROS generation (C. Li, Ge, Yin, Luo, & Chen, 2012). The rs4808209 (in *ZNF1* gene) is an eQTL for *LPAR2* - the receptor for a signaling lipid LPA. It is a part of the LPAR2/Gab1/PI3K/Akt pathway, which influences glucose uptake (Rodriguez-Araujo et al., 2013) and is differentially expressed in fatty vs normal liver in extreme obese cohort (DiStefano et al., 2014). Its expression correlates with the invasiveness of pancreatic cancers (Gong et al., 2012) as well as gynecological disorders (Fujii et al., 2019; Kowalczyk-Zieba et al., 2019; Wasniewski & Woclawek-Potocka, 2018; X. Yu, Zhang, & Chen, 2016).

In Analyses 3 and 4, we set to determine whether genes associated with GWG also impact BMI. Basing on the summary results from the GIANT cohort we show a number of such genes. Some of their SNPs were previously associated with BMI, body fat distribution, attendance to gym or sport groups, sleep duration, height, lipoprotein levels, low HDL-level cholesterol. At the same time, in a much smaller ARIC cohort we find 15 variants associated with diabetes adjusted BMI. None of them is listed in GWAS Catalog or PheWas, however other variants in these genes were already mentioned to be associated with BMI.

Our analysis shows association with rs12685009 in the *SLC24A2* loci, which belongs to the family of K^+^ dependent Na^+^/Ca^2+^ exchanger (Schnetkamp, 2013). A member of the family, SLC4 influences MC4R dependent satiety and the loss of MC4R function leads to prolonged obesity and reduced energy expenditure, while mice lacking *SLC4* display anorexia (X. F. Li & Lytton, 2014). Also, variants in *SOX6*, a transcription factor shown to be involved in the promotion of adipogenesis (Iguchi et al., 2005; Leow et al., 2016), were previously related to obesity (Correa-Rodríguez, Schmidt-RioValle, & Rueda-Medina, 2018; Y. Z. Liu et al., 2009). The rs77346003 in the *ADIPOR1* loci - a receptor for adiponectin known to regulate glucose metabolism and fatty acid oxidation associates with BMI in our study. Its variants in GWAS are related to obesity (Beckers et al., 2013; Keustermans et al., 2017; Peters et al., 2013) as well as fetal weight (Fensterseifer, Austin, Ford, & Alexander, 2018; Muñoz-Muñoz, Krause, Uauy, & Casanello, 2018). ADIPOR1 expression correlates with improved insulin sensitivity and PGC-1A expression - a master regulator of mitochondrial gene expression (Za’don et al., 2019). Moreover, our analysis identifies eQTLs for mitochondria associated genes – *ATP5G3* (rs144768710) and *HSPB8* (rs4131847). ATP5G3 is involved in the proton pathway and acts as an energy-driving motor (He et al., 2017; Huang et al., 2013; Spataru, Le Duc, Zagrean, & Zagrean, 2019). HSPB8 is known to prevent oxidative tissue damage and its expression in serum is used as a biomarker for virus induced type 1 diabetes (Karthik, Ilavenil, Kaleeswaran, Sunil, & Ravikumar, 2012; X. C. Li et al., 2017; L. Yu et al., 2019). We also identified rs34427781 - an eQTL for several genes (*CYP2D8P, NDUFA6-AS1, CYP2D6, CCDC134, CENPM, CHADL, FAM109B, NAGA, NDUFA6, OGFRP1, OLA1P1, SEPT3, SHISA8, SLC25A5P1, SMDT1, SREBF2, TNFRSF13C, WBP2NL*) to be associated with BMI. It is worth noting that a subset of these genes is involved in lipid metabolism (*NAGA, SREBP*) or mitochondrial signaling (*SLC25A5P1, NDUFA6, SMDT1*). SREBP is a master regulator of lipid and sterol biosynthesis (Düvel et al., 2010); the gene-gene interactions between variants in the INSIG-SCAP-SREBP pathway are associated with risk of obesity in Chinese children (F. H. Liu et al., 2014). The *SMDT1* gene builds the mitochondrial calcium uniporter subunit (mtCU) localized in the inner mitochondrial membrane and is responsible for the Ca^2+^ transport to the mitochondrial matrix (Pendin, Greotti, & Pozzan, 2014), while its expression was shown to associate with fetal development (Vishnyakova et al., 2019). *SLC25* genes, members of SLC channels mentioned above, are known to be subunits of another complex essential for proper mitochondrial dynamics and energy production – ANC. ANC is responsible for mitochondrial/cytoplasmic ADP/ATP exchange (Clémençon, Babot, & Trézéguet, 2013) and its expression was shown to be consistently upregulated in obesity (Padilla et al., 2014). NDUFA6 is part of the first complex of the mitochondrial respiratory chain, which expression impacts oxidative stress and energy production efficacy (Fiedorczuk & Sazanov, 2018). Lastly, the *TNFRSF13C* (*BAFF*) gene regulates insulin sensitivity (Kawasaki et al., 2013) and is associated with autoimmune diseases (Moisini & Davidson, 2009).

The above results are congruent with the gene-based fBAT analysis, that points us towards genes which correlate with BMI (*GATAD2, LHFPL6*) and immune response/autoimmunity (*HLA-DQB, HLA-DRB, HLA-B*). GATAD2A is involved in embryonic development (Wang et al., 2017) and associates with obesity (Saxena et al., 2012). A copy number variation of *LHFPL6* associates with average daily gain in cattle (Xu et al., 2019), while its rs4073643 with systemic lupus erythematosus in the Chinese population (Zhang et al., 2016). While the association between HLA variants and T1DM is very well known (Zhao et al., 2016), HLA loci are linked to other phenotypes (Karnes et al., 2017): type 2 diabetes (Ng et al., 2014), fatty liver disease (Doganay et al., 2014) as well as BMI (Shen et al., 2018) or waist-to-hip circumference (Wen et al., 2016). More importantly, these variants associate with trends in fetal birth weight (Capittini et al., 2009), level of inflammation in visceral adipose tissue in pregnant women (Eyerahi et al., 2018) and different mRNA levels in visceral omental adipose tissue of pregnant women with gestational diabetes (Deng et al., 2018). The *HLA-DQA1, HLADQB1* are differentially methylated in siblings born before vs after maternal bariatric surgery (Berglind et al., 2016).

In this study we identified several loci which contribute to the genetic correlation between BMI and GWG. Variants identified in those loci are associated with genes linked to insulin signaling, glucose homeostasis, mitochondrial metabolism, ubiquitinylation and inflammatory responses and placenta functioning, not only in the diabetic cohorts, but also in the general population. The genetic contribution to GWG is clearly connected with BMI associated loci.

## Data Availability

The data are available for reasearch use only. Ethics approval is reguired to access the data.

https://ega-archive.org/studies/EGAS00001004408

## DECLARATIONS

### Ethics approval and consent to participate

This study was approved by the Bioethical Committees of the Jagiellonian University and Poznan University of Medical Sciences and performed according to the Helsinki Declaration.

### Consent to participate

Informed consent was obtained from all individual participants included in the study.

### Consent to publication

Not applicable

### Availability of data and materials

Sequence data has been deposited at the European Genome-phenome Archive (EGA), which is hosted by the EBI and the CRG, under accession number EGAS00001004408. Further information about EGA can be found on https://ega-archive.org “The European Genome-phenome Archive of human data consented for biomedical research” (http://www.nature.com/ng/journal/v47/n7/full/ng.3312.html).

## ACKNOWLEDGEMENTS

The authors thank prof. Katarzyna Cypryk and dr Monika Żurawska-Kliś for their help in recruiting patients.

This research was supported in part by PLGrid Infrastructure. The bioinformatic analysis was performed using Prometheus (AGH, Krakow, Poland), Michigan Imputaton Server (Michigan, MI, USA) and The Ohio Supercomputer Center (Columbus, OH, USA).

For manuscripts with ARIC GWAS data (including HapMap and 1000G imputed data): The Atherosclerosis Risk in Communities study has been funded in whole or in part with Federal funds from the National Heart, Lung, and Blood Institute, National Institutes of Health, Department of Health and Human Services (contract numbers HHSN268201700001I, HHSN268 201700002I, HHSN268201700003I, HHSN268201700004I and HHSN268201700005I), R0 1HL087641, R01HL059367 and R01HL086694; National Human Genome Research Institute contract U01HG004402; and National Institutes of Health contract HHSN268200625226C. The authors thank the staff and participants of the ARIC study for their important contributions. Infrastructure was partly supported by Grant Number UL1RR025005, a component of the National Institutes of Health and NIH Roadmap for Medical Research.

The BioMe Biobank Program T2D-GENES Exome Sequencing Study was conducted by the BioMe Biobank Program T2D-GENES Exome Sequencing Study Investigators and supported by the National Institute of Diabetes and Digestive and Kidney Diseases (NIDDK). The data from the BioMe Biobank Program T2D-GENES Exome Sequencing Study reported here were supplied by the Broad Institute and Icahn School of Medicine at Mount Sinai. This manuscript was not prepared in collaboration with Investigators of the BioMe Biobank Program T2D-GENES Exome Sequencing study and does not necessarily reflect the opinions or views of the BioMe Biobank Program T2D-GENES Exome Sequencing study, or the NIDDK.

## Authorship statement

All authors whose names appear on the submission made substantial contributions to the conception or design of the work; or the acquisition, analysis, or interpretation of data; or the creation of new software used in the work; drafted the work or revised it critically for important intellectual content; approved the version to be published; agree to be accountable for all aspects of the work in ensuring that questions related to the accuracy or integrity of any part of the work are appropriately investigated and resolved.

## SUPPLEMENTARY INFORMATION LEGEND

Table S1a. The results of PrediXcan on BMI in T1D cohort in Subcutaneous Adipose Tissue.

Table S1b. The results of PrediXcan on BMI in T1D cohort in Visceral Adipose Tissue.

Table S1c. The results of PrediXcan on GWG in T1D cohort in Subcutaneous Adipose Tissue.

Table S1d. The results of PrediXcan on GWG in T1D cohort in Visceral Adipose Tissue.

Table S2. The overlap of PrediXcan results in Subcutaneous and Visceral Adipose Tissue in the Giant cohort.

Table 3a. The results of PrediXcan on Giant cohort on BMI only.

Table S3b. The results of PrediXcan on Giant cohort on BMI and GWG.

Table S3c. The results of PrediXcan on Giant cohort on GWG only.

Table S4a.The results of PrediXcan on BMI in T2D cohort in Subcutaneous Adipose Tissue.

Table S4b. The results of PrediXcan on BMI in T2D cohort in Visceral Adipose Tissue.

Table S4c. The results of PrediXcan on BMI in ARIC cohort in Subcutaneous Adipose Tissue.

Table S4d. The results of PrediXcan on BMI in ARIC cohort in Visceral Adipose Tissue.

Table S5a. The results of PrediXcan on T2D&ARIC cohorts on BMI only.

Table S5b. The results of PrediXcan on T2D&ARIC cohorts on BMI and GWG. Table S5c. The results of PrediXcan on T2D&ARIC cohorts on GWG only.

Table S6. The list of variants associated with GWG in BMI associated genes in T2D&ARIC cohorts.

Table S7. The FUMA analysis on BMI associated variants in GWG associated genes in T1DM cohort.

Table S8a. Go Enrichment analysis on the overlap between BMI and GWG genes in the Giant cohort.

Table S8b. Go Enrichment analysis on BMI only associated genes in the Giant cohort. Table S8c. Go Enrichment analysis on GWG only associated genes in the Giant cohort.

Table S9a. Go Enrichment analysis on the overlap between BMI and GWG genes in T2D and ARIC cohorts.

Table s9b. Go Enrichment analysis on BMI only associated genes in T2D and ARIC cohorts.

Table S9c. Go Enrichment analysis on GWG only associated genes in T2D and ARIC cohorts.

Figure S1a. The LD analysis for GPN3 gene. Figure S1b. The LD analysis for PMS2P3 gene.

Figure S1c. The LD analysis for STAG3L1 gene.

Figure S2. The KEGG analysis on GWG associated variants in BMI associated genes in T2D&ARIC cohorts.

Figure S3a. Go Enrichment analysis on GWG only associated genes in the Giant cohort.

Figure S3b. Go Enrichment analysis on the overlap between BMI and GWG genes in the Giant cohort.

Figure S4a. Go Enrichment analysis on GWG only associated genes in T2D and ARIC cohorts.

Figure S4b. Go Enrichment analysis on the overlap between BMI and GWG genes in T2D and ARIC cohorts.

## REFERENCES

Andersson, E. S., Silventoinen, K., Tynelius, P., Nohr, E. A., Sørensen, T. I. A., & Rasmussen, F. (2015). Heritability of Gestational Weight Gain - A Swedish Register-Based Twin Study. Twin Research and Human Genetics. https://doi.org/10.1017/thg.2015.38

Barbati, S. A., Colussi, C., Bacci, L., Aiello, A., Re, A., Stigliano, E., … Nanni, S. (2017). Transcription factor CREM mediates high glucose response in cardiomyocytes and in a male mouse model of prolonged hyperglycemia. Endocrinology. https://doi.org/10.1210/en.2016-1960

Bashir, M., Naem, E., Taha, F., Konje, J. C., & Abou-Samra, A. B. (2019). Outcomes of type 1 diabetes mellitus in pregnancy; effect of excessive gestational weight gain and hyperglycaemia on fetal growth. Diabetes and Metabolic Syndrome: Clinical Research and Reviews. https://doi.org/10.1016/j.dsx.2018.08.030

Beckers, S., De Freitas, F., Zegers, D., Mertens, I. L., Verrijken, A., Van Camp, J. K., … Van Hul, W. (2013). No conclusive evidence for association of polymorphisms in the adiponectin receptor 1 gene, AdipoR1, with common obesity. Endocrine. https://doi.org/10.1007/s12020-012-9736-6

Berglind, D., Müller, P., Willmer, M., Sinha, I., Tynelius, P., Näslund, E., … Rasmussen, F. (2016). Differential methylation in inflammation and type 2 diabetes genes in siblings born before and after maternal bariatric surgery. Obesity. https://doi.org/10.1002/oby.21340

Capittini, C., Pasi, A., Bergamaschi, P., Tinelli, C., De Silvestri, A., Mercati, M. P., … Cuccia, M. (2009). HLA haplotypes and birth weight variation: Is your future going to be light or heavy? Tissue Antigens. https://doi.org/10.1111/j.1399-0039.2009.01282.x

Celia, B., Richard, H., Chineze, O., Casey, E., Elizabeth, P., & Macauley, A. (2016). The St Vincent Declaration–25 years on: review of maternal deaths reported to the Confidential Enquiry in the UK and the lessons learnt. European Journal of Obstetrics & Gynecology and Reproductive Biology. https://doi.org/10.1016/j.ejogrb.2016.07.354

Cheng, M., Mei, B., Zhou, Q., Zhang, M., Huang, H., Han, L., & Huang, Q. (2018). Computational analyses of obesity associated loci generated by genome-wide association studies. PLoS ONE. https://doi.org/10.1371/journal.pone.0199987

Clémençon, B., Babot, M., & Trézéguet, V. (2013). The mitochondrial ADP/ATP carrier (SLC25 family): Pathological implications of its dysfunction. Molecular Aspects of Medicine. https://doi.org/10.1016/j.mam.2012.05.006

Correa-Rodríguez, M., Schmidt-RioValle, J., & Rueda-Medina, B. (2018). SOX6 rs7117858 polymorphism is associated with osteoporosis and obesity-related phenotypes. European Journal of Clinical Investigation. https://doi.org/10.1111/eci.13011

Deng, X., Yang, Y., Sun, H., Qi, W., Duan, Y., & Qian, Y. (2018). Analysis of whole genome-wide methylation and gene expression profiles in visceral omental adipose tissue of pregnancies with gestational diabetes mellitus. Journal of the Chinese Medical Association. https://doi.org/10.1016/j.jcma.2017.06.027

DiStefano, J. K., Kingsley, C., Craig Wood, G., Chu, X., Argyropoulos, G., Still, C. D., … Gerhard, G. S. (2014). Genome-wide analysis of hepatic lipid content in extreme obesity. Acta Diabetologica. https://doi.org/10.1007/s00592-014-0654-3

Doganay, L., Katrinli, S., Colak, Y., Senates, E., Zemheri, E., Ozturk, O., … Doganay, G. D. (2014). HLA DQB1 alleles are related with nonalcoholic fatty liver disease. Molecular Biology Reports. https://doi.org/10.1007/s11033-014-3688-2

Dori-Dayan, N., Cukierman-Yaffe, T., Zemet, R., Cohen, O., Mazaki-Tovi, S., & Yoeli-Ullman, R. (2020). 1013: Gestational weight gain does-not affect insulin requirement during pregnancy in women with Type 1 diabetes. American Journal of Obstetrics and Gynecology. https://doi.org/10.1016/j.ajog.2019.11.1029

Düvel, K., Yecies, J. L., Menon, S., Raman, P., Lipovsky, A. I., Souza, A. L., … Manning, B. D. (2010). Activation of a metabolic gene regulatory network downstream of mTOR complex 1. Molecular Cell. https://doi.org/10.1016/j.molcel.2010.06.022

Ellis, K. L., Zhou, Y., Beshansky, J. R., Ainehsazan, E., Selker, H. P., Cupples, L. A., … Peter, I. (2015). Genetic modifiers of response to glucose-insulin-potassium (GIK) infusion in acute coronary syndromes and associations with clinical outcomes in the IMMEDIATE trial. Pharmacogenomics Journal. https://doi.org/10.1038/tpj.2015.10

Eyerahi, B. F., Mancilla-Herrera, I., Espino Y Sosa, S., Ortiz-Ramirez, M., Flores-Rueda, V., Ibargüengoitia-Ochoa, F., … Estrada-Gutierrez, G. (2018). Macrophage populations in visceral adipose tissue from pregnant women: Potential role of obesity in maternal inflammation. International Journal of Molecular Sciences. https://doi.org/10.3390/ijms19041074

Fang, C. Y., Yu, C. C., Liao, Y. W., Hsieh, P. L., Lu, M. Y., Lin, K. C., … Tsai, L. L. (2019). LncRNA LINC00974 activates TGF-β/Smad signaling to promote oral fibrogenesis. Journal of Oral Pathology and Medicine. https://doi.org/10.1111/jop.12805

Fensterseifer, S. R., Austin, K. J., Ford, S. P., & Alexander, B. M. (2018). Effects of maternal obesity on maternal and fetal plasma concentrations of adiponectin and expression of adiponectin and its receptor genes in cotyledonary and adipose tissues at mid- and late-gestation in sheep. Animal Reproduction Science. https://doi.org/10.1016/j.anireprosci.2018.08.033

Fiedorczuk, K., & Sazanov, L. A. (2018). Mammalian Mitochondrial Complex I Structure and Disease-Causing Mutations. Trends in Cell Biology. https://doi.org/10.1016/j.tcb.2018.06.006

Finlin, B. S., Zhu, B., Confides, A. L., Westgate, P. M., Harfmann, B. D., Dupont-Versteegden, E. E., & Kern, P. A. (2017). Mast cells promote seasonal white adipose beiging in humans. Diabetes. https://doi.org/10.2337/db16-1057

Forde, N., Duffy, G. B., McGettigan, P. A., Browne, J. A., Mehta, J. P., Kelly, A. K., … Evans, A. C. O. (2012). Evidence for an early endometrial response to pregnancy in cattle: Both dependent upon and independent of interferon tau. Physiological Genomics. https://doi.org/10.1152/physiolgenomics.00067.2012

Fujii, T., Nagamatsu, T., Schust, D. J., Ichikawa, M., Kumasawa, K., Yabe, S., … Fujii, T. (2019). Placental expression of lysophosphatidic acid receptors in normal pregnancy and preeclampsia. American Journal of Reproductive Immunology. https://doi.org/10.1111/aji.13176

Gamazon, E. R., Wheeler, H. E., Shah, K. P., Mozaffari, S. V., Aquino-Michaels, K., Carroll, R. J., … Im, H. K. (2015). A gene-based association method for mapping traits using reference transcriptome data. Nature Genetics. https://doi.org/10.1038/ng.3367

Gentili, A., Zaibi, M. S., Alomar, S. Y., De Vuono, S., Ricci, M. A., Alaeddin, A., … Lupattelli, G. (2016). Circulating Levels of the Adipokines Monocyte Chemotactic Protein-4 (MCP-4), Macrophage Inflammatory Protein-1β (MIP-1β), and Eotaxin-3 in Severe Obesity and Following Bariatric Surgery. Hormone and Metabolic Research. https://doi.org/10.1055/s-0042-108731

Gong, Y. L., Tao, C. J., Hu, M., Chen, J. F., Cao, X. F., Lv, G. M., & Li, P. (2012). Expression of lysophosphatidic acid receptors and local invasiveness and metastasis in Chinese pancreatic cancers. Current Oncology. https://doi.org/10.3747/co.19.1138

Han, K., Hassanzadeh, S., Singh, K., Menazza, S., Nguyen, T. T., Stevens, M. V., … Sack, M. N. (2017). Parkin regulation of CHOP modulates susceptibility to cardiac endoplasmic reticulum stress. Scientific Reports. https://doi.org/10.1038/s41598-017-02339-2

He, J., Ford, H. C., Carroll, J., Ding, S., Fearnley, I. M., & Walker, J. E. (2017). Persistence of the mitochondrial permeability transition in the absence of subunit c of human ATP synthase. Proceedings of the National Academy of Sciences of the United States of America. https://doi.org/10.1073/pnas.1702357114

Hebbar, P., Abu-Farha, M., Alkayal, F., Nizam, R., Elkum, N., Melhem, M., … Thanaraj, T. A. (2020). Genome-wide association study identifies novel risk variants from RPS6KA1, CADPS, VARS, and DHX58 for fasting plasma glucose in Arab population. Scientific Reports. https://doi.org/10.1038/s41598-019-57072-9

Hormozdiari, F., van de Bunt, M., Segrè, A. V., Li, X., Joo, J. W. J., Bilow, M., … Eskin, E. (2016). Colocalization of GWAS and eQTL Signals Detects Target Genes. American Journal of Human Genetics. https://doi.org/10.1016/j.ajhg.2016.10.003

Hsiung, C.-N., Chang, Y.-C., Lin, C.-W., Chang, C.-W., Chou, W.-C., Chu, H.-W., … Shen, C.-Y. (2019). The Causal Relationship of Circulating Triglycerides and Glycated Hemoglobin: A Mendelian Randomization Study. The Journal of Clinical Endocrinology & Metabolism. https://doi.org/10.1210/clinem/dgz243

Hu, Y. W., Guo, F. X., Xu, Y. J., Li, P., Lu, Z. F., McVey, D. G., … Ye, S. (2019). Long noncoding RNA NEXN-AS1 mitigates atherosclerosis by regulating the actin-binding protein NEXN. Journal of Clinical Investigation. https://doi.org/10.1172/JCI98230

Huang, Y., Wang, L., Bennett, B., Williams, R. W., Wang, Y. J., Gu, W. K., & Jiao, Y. (2013). Potential role of Atp5g3 in epigenetic regulation of alcohol preference or obesity from a mouse genomic perspective. Genetics and Molecular Research. https://doi.org/10.4238/2013.September.18.1

Huyghe, J. R., Jackson, A. U., Fogarty, M. P., Buchkovich, M. L., Stančáková, A., Stringham, H. M., … Mohlke, K. L. (2013). Exome array analysis identifies new loci and low-frequency variants influencing insulin processing and secretion. Nature Genetics. https://doi.org/10.1038/ng.2507

Hwang, J. Y., Sim, X., Wu, Y., Liang, J., Tabara, Y., Hu, C., … Kim, B. J. (2015). Genome-wide association meta-analysis identifies novel variants associated with fasting plasma glucose in East Asians. Diabetes. https://doi.org/10.2337/db14-0563

Ignacio, R. M. C., Gibbs, C. R., Lee, E. S., & Son, D. S. (2016). Differential chemokine signature between human preadipocytes and adipocytes. Immune Network. https://doi.org/10.4110/in.2016.16.3.189

Iguchi, H., Ikeda, Y., Okamura, M., Tanaka, T., Urashima, Y., Ohguchi, H., … Sakai, J. (2005). SOX6 attenuates glucose-stimulated insulin secretion by repressing PDX1 transcriptional activity and is down-regulated in hyperinsulinemic obese mice. Journal of Biological Chemistry. https://doi.org/10.1074/jbc.M505392200

Influence of Pregnancy Weight on Maternal and Child Health. (2007). Influence of Pregnancy Weight on Maternal and Child Health. https://doi.org/10.17226/11817

Kang, M., Lee, S. M., Kim, W., Lee, K. H., & Kim, D. Y. (2019). Fubp1 supports the lactate-Akt-mTOR axis through the upregulation of Hk1 and Hk2. Biochemical and Biophysical Research Communications. https://doi.org/10.1016/j.bbrc.2019.03.005

Karnes, J. H., Bastarache, L., Shaffer, C. M., Gaudieri, S., Xu, Y., Glazer, A. M., … Denny, J. C. (2017). Phenome-wide scanning identifies multiple diseases and disease severity phenotypes associated with HLA variants. Science Translational Medicine. https://doi.org/10.1126/scitranslmed.aai8708

Karthik, D., Ilavenil, S., Kaleeswaran, B., Sunil, S., & Ravikumar, S. (2012). Proteomic analysis of plasma proteins in diabetic rats by 2D electrophoresis and MALDI-TOF-MS. Applied Biochemistry and Biotechnology. https://doi.org/10.1007/s12010-012-9544-8

Katoh, M. (2013). Functional proteomics, human genetics and cancer biology of GIPC family members. Experimental and Molecular Medicine. https://doi.org/10.1038/emm.2013.49

Kawai, V. K., Nwosu, S. K., Kurnik, D., Harrell, F. E., & Stein, C. M. (2019). Variants in BMI-Associated Genes and Adrenergic Genes are not Associated with Gestational Weight Trajectory. Obesity. https://doi.org/10.1002/oby.22505

Kawasaki, K., Abe, M., Tada, F., Tokumoto, Y., Chen, S., Miyake, T., … Onji, M. (2013). Blockade of B-cell-activating factor signaling enhances hepatic steatosis induced by a high-fat diet and improves insulin sensitivity. Laboratory Investigation. https://doi.org/10.1038/labinvest.2012.176

Kershaw, E. E., Hamm, J. K., Verhagen, L. A. W., Peroni, O., Katic, M., & Flier, J. S. (2006). Adipose triglyceride lipase: Function, regulation by insulin, and comparison with adiponutrin. Diabetes. https://doi.org/10.2337/diabetes.55.01.06.db05-0982

Keustermans, G., Van Der Heijden, L. B., Boer, B., Scholman, R., Nuboer, R., Pasterkamp, G., … Schipper, H. S. (2017). Differential adipokine receptor expression on circulating leukocyte subsets in lean and obese children. PLoS ONE. https://doi.org/10.1371/journal.pone.0187068

Kobayashi, H., Yamazaki, S., Takashima, S., Liu, W., Okuda, H., Yan, J., … Koizumi, A. (2013). Ablation of Rnf213 retards progression of diabetes in the Akita mouse. Biochemical and Biophysical Research Communications. https://doi.org/10.1016/j.bbrc.2013.02.015

Kominiarek, M. A., & Peaceman, A. M. (2017). Gestational weight gain. American Journal of Obstetrics and Gynecology. https://doi.org/10.1016/j.ajog.2017.05.040

Kowalczyk-Zieba, I., Woclawek-Potocka, I., Wasniewski, T., Boruszewska, D., Grycmacher, K., Sinderewicz, E., … Wolczynski, S. (2019). LPAR2 and LPAR4 are the Main Receptors Responsible for LPA Actions in Ovarian Endometriotic Cysts. Reproductive Sciences. https://doi.org/10.1177/1933719118766263

Kusić, D., Connolly, J., Kainulainen, H., Semenova, E. A., Borisov, O. V., Larin, A. K., … Burniston, J. G. (2020). Striated muscle-specific serine/threonine-protein kinase beta segregates with high versus low responsiveness to endurance exercise training. Physiological Genomics. https://doi.org/10.1152/physiolgenomics.00103.2019

Lawlor, D. A., Fraser, A., Macdonald-Wallis, C., Nelson, S. M., Palmer, T. M., Smith, G. D., & Tilling, K. (2011). Maternal and offspring adiposity-related genetic variants and gestational weight gain. American Journal of Clinical Nutrition. https://doi.org/10.3945/ajcn.110.010751

Leow, S. C., Poschmann, J., Too, P. G., Yin, J., Joseph, R., McFarlane, C., … Stünkel, W. (2016). The transcription factor SOX6 contributes to the developmental origins of obesity by promoting adipogenesis. Development (Cambridge). https://doi.org/10.1242/dev.131573

Li, B., Verma, S. S., Veturi, Y. C., Verma, A., Bradford, Y., Haas, D. W., & Ritchie, M. D. (2018). Evaluation of predixcan for prioritizing GWAS associations and predicting gene expression. In Pacific Symposium on Biocomputing. https://doi.org/10.1142/9789813235533_0041

Li, C., Ge, M., Yin, Y., Luo, M., & Chen, D. (2012). Silencing expression of ribosomal protein L26 and L29 by RNA interfering inhibits proliferation of human pancreatic cancer PANC-1 cells. Molecular and Cellular Biochemistry. https://doi.org/10.1007/s11010-012-1404-x

Li, X. C., Hu, Q. K., Chen, L., Liu, S. Y., Su, S., Tao, H., … He, L. J. (2017). HSPB8 promotes the fusion of autophagosome and lysosome during autophagy in diabetic neurons. International Journal of Medical Sciences. https://doi.org/10.7150/ijms.20653

Li, X. F., & Lytton, J. (2014). An essential role for the K+-dependent Na+/Ca2+-exchanger, NCKX4, in melanocortin-4-receptor-dependent satiety. Journal of Biological Chemistry. https://doi.org/10.1074/jbc.M114.564450

Lin, H. M., Lee, J. H., Yadav, H., Kamaraju, A. K., Liu, E., Zhigang, D., … Rane, S. G. (2009). Transforming growth factor-β/Smad3 signaling regulates insulin gene transcription and pancreatic islet β-cell function. Journal of Biological Chemistry. https://doi.org/10.1074/jbc.M805379200

Liu, F. H., Song, J. Y., Shang, X. R., Meng, X. R., Ma, J., & Wang, H. J. (2014). The gene-gene interaction of INSIG-SCAP-SREBP pathway on the risk of obesity in Chinese children. BioMed Research International. https://doi.org/10.1155/2014/538564

Liu, Y. Z., Pei, Y. F., Liu, J. F., Yang, F., Guo, Y., Zhang, L., … Deng, H. W. (2009). Powerful bivariate Genome-wide association analyses suggest the SOX6 gene influencing both obesity and osteoporosis phenotypes in males. PLoS ONE. https://doi.org/10.1371/journal.pone.0006827

Liu, Z., Li, C., Kang, N., Malhi, H., Shah, V. H., & Maiers, J. L. (2019). Transforming growth factor (TGF) cross-talk with the unfolded protein response is critical for hepatic stellate cell activation. Journal of Biological Chemistry. https://doi.org/10.1074/jbc.RA118.005761

Locke, A., Kahali, B., Berndt, S., Justice, A., & Pers, T. (2015). Genetic studies of body mass index yield new insights for obesity biology. Nature, 518(7538), 197–206. https://doi.org/10.1038/nature14177.Genetic

Loucks, C. M., Bialas, N. J., Dekkers, M. P. J., Walker, D. S., Grundy, L. J., Li, C., … Leroux, M. R. (2016). PACRG, a protein linked to ciliary motility, mediates cellular signaling. Molecular Biology of the Cell. https://doi.org/10.1091/mbc.E15-07-0490

Ludwig-Slomczynska, A. H., Seweryn, M. T., Kapusta, P., Pitera, E., Mantaj, U., Cyganek, K., … Wolkow, P. P. (2018). Mitochondrial GWAS and association of nuclear-mitochondrial epistasis with BMI in T1DM patients. BioRxiv. https://doi.org/10.1101/436519

Luecke, E., Cohen, A. K., Brillante, M., Rehkopf, D. H., Coyle, J., Hendrick, C. E., & Abrams, B. (2018). Similarities in Maternal Weight and Birth Weight Across Pregnancies and Across Sisters. Maternal and Child Health Journal. https://doi.org/10.1007/s10995-018-2602-2

Mancuso, N., Shi, H., Goddard, P., Kichaev, G., Gusev, A., & Pasaniuc, B. (2017). Integrating Gene Expression with Summary Association Statistics to Identify Genes Associated with 30 Complex Traits. American Journal of Human Genetics. https://doi.org/10.1016/j.ajhg.2017.01.031

Mastella, L. S., Weinert, L. S., Gnielka, V., Hirakata, V. N., Oppermann, M. L. R., Silveiro, S. P., & Reichelt, A. J. (2018). Influence of maternal weight gain on birth weight: A gestational diabetes cohort. Archives of Endocrinology and Metabolism. https://doi.org/10.20945/2359-3997000000009

McWhorter, K. L., Bowers, K., Dolan, L. M., Deka, R., Jackson, C. L., & Khoury, J. C. (2018). Impact of gestational weight gain and prepregnancy body mass index on the prevalence of large-for-gestational age infants in two cohorts of women with type 1 insulin-dependent diabetes: A cross-sectional population study. BMJ Open. https://doi.org/10.1136/bmjopen-2017-019617

Meschede, J., Šadić, M., Furthmann, N., Miedema, T., Sehr, D. A., Müller-Rischart, A. K., … Winklhofer, K. F. (2020). The parkin-coregulated gene product PACRG promotes TNF signaling by stabilizing LUBAC. Science Signaling. https://doi.org/10.1126/scisignal.aav1256

Moisini, I., & Davidson, A. (2009). BAFF: A local and systemic target in autoimmune diseases. Clinical and Experimental Immunology. https://doi.org/10.1111/j.1365-2249.2009.04007.x

Muñoz-Muñoz, E., Krause, B. J., Uauy, R., & Casanello, P. (2018). LGA-newborn from patients with pregestational obesity present reduced adiponectin-mediated vascular relaxation and endothelial dysfunction in fetoplacental arteries. Journal of Cellular Physiology. https://doi.org/10.1002/jcp.26499

Muro, S., Miyake, Y., Kato, H., Tsutsumi, K., & Yamamoto, K. (2015). Serum anti-60s ribosomal protein L29 antibody as a novel prognostic marker for unresectable pancreatic cancer. Digestion. https://doi.org/10.1159/000371545

Ng, M. C. Y., Shriner, D., Chen, B. H., Li, J., Chen, W. M., Guo, X., … Surdulescu, G. (2014). Meta-Analysis of Genome-Wide Association Studies in African Americans Provides Insights into the Genetic Architecture of Type 2 Diabetes. PLoS Genetics. https://doi.org/10.1371/journal.pgen.1004517

Nunnery, D., Ammerman, A., & Dharod, J. (2018). Predictors and outcomes of excess gestational weight gain among low-income pregnant women. Health Care for Women International. https://doi.org/10.1080/07399332.2017.1391263

Padilla, J., Jenkins, N. T., Thorne, P. K., Martin, J. S., Scott Rector, R., Wade Davis, J., & Harold Laughlin, M. (2014). Identification of genes whose expression is altered by obesity throughout the arterial tree. Physiological Genomics. https://doi.org/10.1152/physiolgenomics.00091.2014

Pendin, D., Greotti, E., & Pozzan, T. (2014). The elusive importance of being a mitochondrial Ca2+ uniporter. Cell Calcium. https://doi.org/10.1016/j.ceca.2014.02.008

Peters, K. E., Beilby, J., Cadby, G., Warrington, N. M., Bruce, D. G., Davis, W. A., … Hung, J. (2013). A comprehensive investigation of variants in genes encoding adiponectin (ADIPOQ) and its receptors (ADIPOR1/R2), and their association with serum adiponectin, type 2 diabetes, insulin resistance and the metabolic syndrome. BMC Medical Genetics. https://doi.org/10.1186/1471-2350-14-15

Piccolis, M., Bond, L. M., Kampmann, M., Pulimeno, P., Chitraju, C., Jayson, C. B. K., … Farese, R. V. (2019). Probing the Global Cellular Responses to Lipotoxicity Caused by Saturated Fatty Acids. Molecular Cell. https://doi.org/10.1016/j.molcel.2019.01.036

Rodriguez-Araujo, G., Nakagami, H., Hayashi, H., Mori, M., Shiuchi, T., Minokoshi, Y., … Kaneda, Y. (2013). Alpha-synuclein elicits glucose uptake and utilization in adipocytes through the Gab1/PI3K/Akt transduction pathway. Cellular and Molecular Life Sciences. https://doi.org/10.1007/s00018-012-1198-8

Rudkowska, I., Pérusse, L., Bellis, C., Blangero, J., Després, J. P., Bouchard, C., & Vohl, M. C. (2015). Interaction between Common Genetic Variants and Total Fat Intake on Low-Density Lipoprotein Peak Particle Diameter: A Genome-Wide Association Study. Journal of Nutrigenetics and Nutrigenomics. https://doi.org/10.1159/000431151

Rys, P. M., Ludwig-Slomczynska, A. H., Cyganek, K., & Malecki, M. T. (2018). Continuous subcutaneous insulin infusion vs multiple daily injections in pregnant women with type 1 diabetes mellitus: A systematic review and meta-analysis of randomised controlled trials and observational studies. European Journal of Endocrinology. https://doi.org/10.1530/EJE-17-0804

Saxena, R., Elbers, C. C., Guo, Y., Peter, I., Gaunt, T. R., Mega, J. L., … Keating, B. J. (2012). Large-scale gene-centric meta-analysis across 39 studies identifies type 2 diabetes loci. American Journal of Human Genetics. https://doi.org/10.1016/j.ajhg.2011.12.022

Schnetkamp, P. P. M. (2013). The SLC24 gene family of Na+/Ca2+-K+ exchangers: From sight and smell to memory consolidation and skin pigmentation. Molecular Aspects of Medicine. https://doi.org/10.1016/j.mam.2012.07.008

Scifres, C. M., Feghali, M. N., Althouse, A. D., Caritis, S. N., & Catov, J. M. (2014). Effect of excess gestational weight gain on pregnancy outcomes in women with type 1 diabetes. Obstetrics and Gynecology. https://doi.org/10.1097/AOG.0000000000000271

Secher, A. L., Parellada, C. B., Ringholm, L., Ásbjörnsdóttir, B., Damm, P., & Mathiesen, E. R. (2014). Higher gestational weight gain is associated with increasing offspring birth weight independent of maternal glycemic control in women with type 1 diabetes. Diabetes Care. https://doi.org/10.2337/dc14-0896

Shen, J., Guo, T., Wang, T., Zhen, Y., Ma, X., Wang, Y., … Baier, L. J. (2018). HLA-B_∗_07, HLA-DRB1_∗_07, HLA-DRB1_∗_12, and HLA-c_∗_03:02 strongly associate with BMI: Data from 1.3 million healthy Chinese adults. Diabetes. https://doi.org/10.2337/db17-0852

Siega-Riz, A. M., Bodnar, L. M., Stotland, N. E., & Stang, J. (2020). The Current Understanding of Gestational Weight Gain Among Women with Obesity and the Need for Future Research. NAM Perspectives. https://doi.org/10.31478/202001a

Song, N. J., Kim, S., Jang, B. H., Chang, S. H., Yun, U. J., Park, K. M., … Park, K. W. (2016). Small molecule-induced complement factor D (Adipsin) promotes lipid accumulation and adipocyte differentiation. PLoS ONE. https://doi.org/10.1371/journal.pone.0162228

Spataru, A., Le Duc, D., Zagrean, L., & Zagrean, A. M. (2019). Ethanol exposed maturing rat cerebellar granule cells show impaired energy metabolism and increased cell death after oxygen-glucose deprivation. Neural Regeneration Research. https://doi.org/10.4103/1673-5374.245474

Speidel, D., Salehi, A., Obermueller, S., Lundquist, I., Brose, N., Renström, E., & Rorsman, P. (2008). CAPS1 and CAPS2 Regulate Stability and Recruitment of Insulin Granules in Mouse Pancreatic β Cells. Cell Metabolism. https://doi.org/10.1016/j.cmet.2007.11.009

Stephenson, S. E. M., Aumann, T. D., Taylor, J. M., Riseley, J. R., Li, R., Mann, J. R., … Lockhart, P. J. (2018). Generation and characterisation of a parkin-Pacrg knockout mouse line and a Pacrg knockout mouse line. Scientific Reports. https://doi.org/10.1038/s41598-018-25766-1

Sugihara, M., Morito, D., Ainuki, S., Hirano, Y., Ogino, K., Kitamura, A., … Nagata, K. (2019). The AAA+ ATPase/ubiquitin ligase mysterin stabilizes cytoplasmic lipid droplets. Journal of Cell Biology. https://doi.org/10.1083/jcb.201712120

Suzuki, T., Gao, J., Ishigaki, Y., Kondo, K., Sawada, S., Izumi, T., … Katagiri, H. (2017). ER Stress Protein CHOP Mediates Insulin Resistance by Modulating Adipose Tissue Macrophage Polarity. Cell Reports. https://doi.org/10.1016/j.celrep.2017.01.076

Tang, J., Zhuo, H., Zhang, X., Jiang, R., Ji, J., Deng, L., … Sun, B. (2014). A novel biomarker Linc00974 interacting with KRT19 promotes proliferation and metastasis in hepatocellular carcinoma. Cell Death and Disease. https://doi.org/10.1038/cddis.2014.518

Vishnyakova, P. A., Tarasova, N. V., Volodina, M. A., Tsvirkun, D. V., Sukhanova, I. A., Kurchakova, T. A., … Vysokikh, M. Y. (2019). Gestation age-associated dynamics of mitochondrial calcium uniporter subunits expression in feto-maternal complex at term and preterm delivery. Scientific Reports. https://doi.org/10.1038/s41598-019-41996-3

Voerman, E., Santos, S., Inskip, H., Amiano, P., Barros, H., Charles, M. A., … Gaillard, R. (2019). Association of Gestational Weight Gain With Adverse Maternal and Infant Outcomes. JAMA. https://doi.org/10.1001/jama.2019.3820

Wang, Z., Kang, J., Deng, X., Guo, B., Wu, B., & Fan, Y. (2017). Knockdown of GATAD2A suppresses cell proliferation in thyroid cancer in vitro. Oncology Reports. https://doi.org/10.3892/or.2017.5436

Warrington, N. M., Richmond, R., Fenstra, B., Myhre, R., Gaillard, R., Paternoster, L., … Lawlor, D. A. (2018). Maternal and fetal genetic contribution to gestational weight gain. International Journal of Obesity. https://doi.org/10.1038/ijo.2017.248

Wasniewski, T., & Woclawek-Potocka, I. (2018). Altered expression of lysophosphatidic acid receptors, in association with the synthesis of estrogens and androgens in type 1 endometrial cancer biology. Gynecological Endocrinology. https://doi.org/10.1080/09513590.2017.1409707

Wen, W., Kato, N., Hwang, J. Y., Guo, X., Tabara, Y., Li, H., … Shu, X. O. (2016). Genome- wide association studies in East Asians identify new loci for waist-hip ratio and waist circumference. Scientific Reports. https://doi.org/10.1038/srep17958

Xu, L., Yang, L., Wang, L., Zhu, B., Chen, Y., Gao, H., … Li, J. (2019). Probe-based association analysis identifies several deletions associated with average daily gain in beef cattle. BMC Genomics. https://doi.org/10.1186/s12864-018-5403-5

Yu, L., Liang, Q., Zhang, W., Liao, M., Wen, M., Zhan, B., … Cheng, X. (2019). HSP22 suppresses diabetes-induced endothelial injury by inhibiting mitochondrial reactive oxygen species formation. Redox Biology. https://doi.org/10.1016/j.redox.2018.101095

Yu, X., Zhang, Y., & Chen, H. (2016). LPA receptor 1 mediates LPA-induced ovarian cancer metastasis: An in vitro and in vivo study. BMC Cancer. https://doi.org/10.1186/s12885-016-2865-1

Za’don, N. H. A., Kamal, A. F. M., Ismail, F., Othman, S. I. T., Appukutty, M., Salim, N., … Ludin, A. F. M. (2019). High-intensity interval training induced PGC-1α and Adipor1 gene expressions and improved insulin sensitivity in obese individuals. Medical Journal of Malaysia.

Zhang, Y., Yang, J., Zhang, J., Sun, L., Hirankarn, N., Pan, H. F., … Yang, W. (2016). Genome-wide search followed by replication reveals genetic interaction of CD80 and ALOX5AP associated with systemic lupus erythematosus in asian populations. Annals of the Rheumatic Diseases. https://doi.org/10.1136/annrheumdis-2014-206367

Zhao, L. P., Alshiekh, S., Zhao, M., Carlsson, A., Larsson, H. E., Forsander, G., … Fureman, A. L. (2016). Next-generation sequencing reveals that HLA-DRB3, -DRB4, and -DRB5 may be associated with islet autoantibodies and risk for childhood type 1 diabetes. Diabetes. https://doi.org/10.2337/db15-1115

Zhu, B., Rippe, C., Holmberg, J., Zeng, S., Perisic, L., Albinsson, S., … Swärd, K. (2018). Nexilin/NEXN controls actin polymerization in smooth muscle and is regulated by myocardin family coactivators and YAP. Scientific Reports. https://doi.org/10.1038/s41598-018-31328-2

